# The *ABO* blood group locus and a chromosome 3 gene cluster associate with SARS-CoV-2 respiratory failure in an Italian-Spanish genome-wide association analysis

**DOI:** 10.1101/2020.05.31.20114991

**Authors:** David Ellinghaus, Frauke Degenhardt, Luis Bujanda, Maria Buti, Agustín Albillos, Pietro Invernizzi, Javier Fernández, Daniele Prati, Guido Baselli, Rosanna Asselta, Marit M Grimsrud, Chiara Milani, Fátima Aziz, Jan Kässens, Sandra May, Mareike Wendorff, Lars Wienbrandt, Florian Uellendahl-Werth, Tenghao Zheng, Xiaoli Yi, Raúl de Pablo, Adolfo Garrido Chercoles, Graduate in Chemistry, Adriana Palom, Alba-Estela Garcia-Fernandez, Francisco Rodriguez-Frias, Alberto Zanella, Alessandra Bandera, Alessandro Protti, Alessio Aghemo, Ana Lleo, Andrea Biondi, Andrea Caballero-Garralda, Andrea Gori, Anja Tanck, Anna Latiano, Anna Ludovica Fracanzani, Anna Peschuk, Antonio Julià, Antonio Pesenti, Antonio Voza, David Jiménez, Beatriz Mateos, Beatriz Nafria Jimenez, Graduate in Biotechnology, Carmen Quereda, Claudio Angelini, Cristina Cea, Aurora Solier, David Pestaña, Elena Sandoval, Elvezia Maria Paraboschi, Enrique Navas, Ferruccio Ceriotti, Filippo Martinelli-Boneschi, Flora Peyvandi, Francesco Blasi, Luis Téllez, Albert Blanco-Grau, Giacomo Grasselli, Giorgio Costantino, Giulia Cardamone, Giuseppe Foti, Serena Aneli, Hayato Kurihara, Hesham ElAbd, Ilaria My, Javier Martin, Jeanette Erdmann, José Ferrusquía-Acosta, Koldo Garcia-Etxebarria, Laura Izquierdo-Sanchez, Graduate in Biochemistry, MS, Laura Rachele Bettini, Leonardo Terranova, Leticia Moreira, Luigi Santoro, Luigia Scudeller, Francisco Mesonero, Luisa Roade, Marco Schaefer, Maria Carrabba, Maria del Mar Riveiro Barciela, Maria Eloina Figuera Basso, Maria Grazia Valsecchi, María Hernandez-Tejero, Marialbert Acosta-Herrera, Mariella D’Angiò, Marina Baldini, Marina Cazzaniga, Martin Schulzky, Maurizio Cecconi, Michael Wittig, Michele Ciccarelli, Miguel Rodríguez-Gandía, Monica Bocciolone, Monica Miozzo, Nicole Braun, Nilda Martínez, Orazio Palmieri, Paola Faverio, Paoletta Preatoni, Paolo Bonfanti, Paolo Omodei, Paolo Tentorio, Pedro Castro, Pedro M. Rodrigues, Aaron Blandino Ortiz, Ricardo Ferrer Roca, Roberta Gualtierotti, Rosa Nieto, Salvatore Badalamenti, Sara Marsal, Giuseppe Matullo, Serena Pelusi, Valter Monzani, Tanja Wesse, Tomas Pumarola, Valeria Rimoldi, Silvano Bosari, Wolfgang Albrecht, Wolfgang Peter, Manuel Romero Gómez, Mauro D’Amato, Stefano Duga, Jesus M. Banales, Johannes R Hov, Trine Folseraas, Luca Valenti, Andre Franke, Tom H Karlsen

## Abstract

**Background:** Respiratory failure is a key feature of severe Covid-19 and a critical driver of mortality, but for reasons poorly defined affects less than 10% of SARS-CoV-2 infected patients.

**Methods:** We included 1,980 patients with Covid-19 respiratory failure at seven centers in the Italian and Spanish epicenters of the SARS-CoV-2 pandemic in Europe (Milan, Monza, Madrid, San Sebastian and Barcelona) for a genome-wide association analysis. After quality control and exclusion of population outliers, 835 patients and 1,255 population-derived controls from Italy, and 775 patients and 950 controls from Spain were included in the final analysis. In total we analyzed 8,582,968 single-nucleotide polymorphisms (SNPs) and conducted a meta-analysis of both case-control panels.

**Results:** We detected cross-replicating associations with rs11385942 at chromosome 3p21.31 and rs657152 at 9q34, which were genome-wide significant (P<5×10^−8^) in the meta-analysis of both study panels, odds ratio [OR], 1.77; 95% confidence interval [CI], 1.48 to 2.11; P=1.14×10^−10^ and OR 1.32 (95% CI, 1.20 to 1.47; P=4.95×10^−8^), respectively. Among six genes at 3p21.31, *SLC6A20* encodes a known interaction partner with angiotensin converting enzyme 2 (ACE2). The association signal at 9q34 was located at the *ABO* blood group locus and a blood-group-specific analysis showed higher risk for A-positive individuals (OR=1.45, 95% CI, 1.20 to 1.75, P=1.48×10^−4^) and a protective effect for blood group O (OR=0.65, 95% CI, 0.53 to 0.79, P=1.06×10^−5^).

**Conclusions:** We herein report the first robust genetic susceptibility loci for the development of respiratory failure in Covid-19. Identified variants may help guide targeted exploration of severe Covid-19 pathophysiology.

## Introduction

Severe acute respiratory syndrome coronavirus 2 (SARS-CoV-2) was discovered in Wuhan in China late 2019 and rapidly evolved into a global pandemic.^1^ As of May 28^th^ 2020, there are over 5.1 million confirmed cases worldwide, with total deaths exceeding 355,000 (access John Hopkins). In Europe, Italy and Spain were early severely affected with epidemic peaks starting in the second half of February 2020 (**Figure 1**) with 60,189 fatal cases reported by May 28^th^ 2020. Coronavirus disease 2019 (Covid-19) has variable behavior,^2^ with the vast majority of infected individuals experiencing only mild or even no symptoms.^3^ Mortality rates are predominantly driven by the subset of patients developing severe respiratory failure secondary to bilateral interstitial pneumonia and acute respiratory distress syndrome.^4^ Severe Covid-19 with respiratory failure requires early and prolonged support by mechanical ventilation.^5^

**Figure 1.**
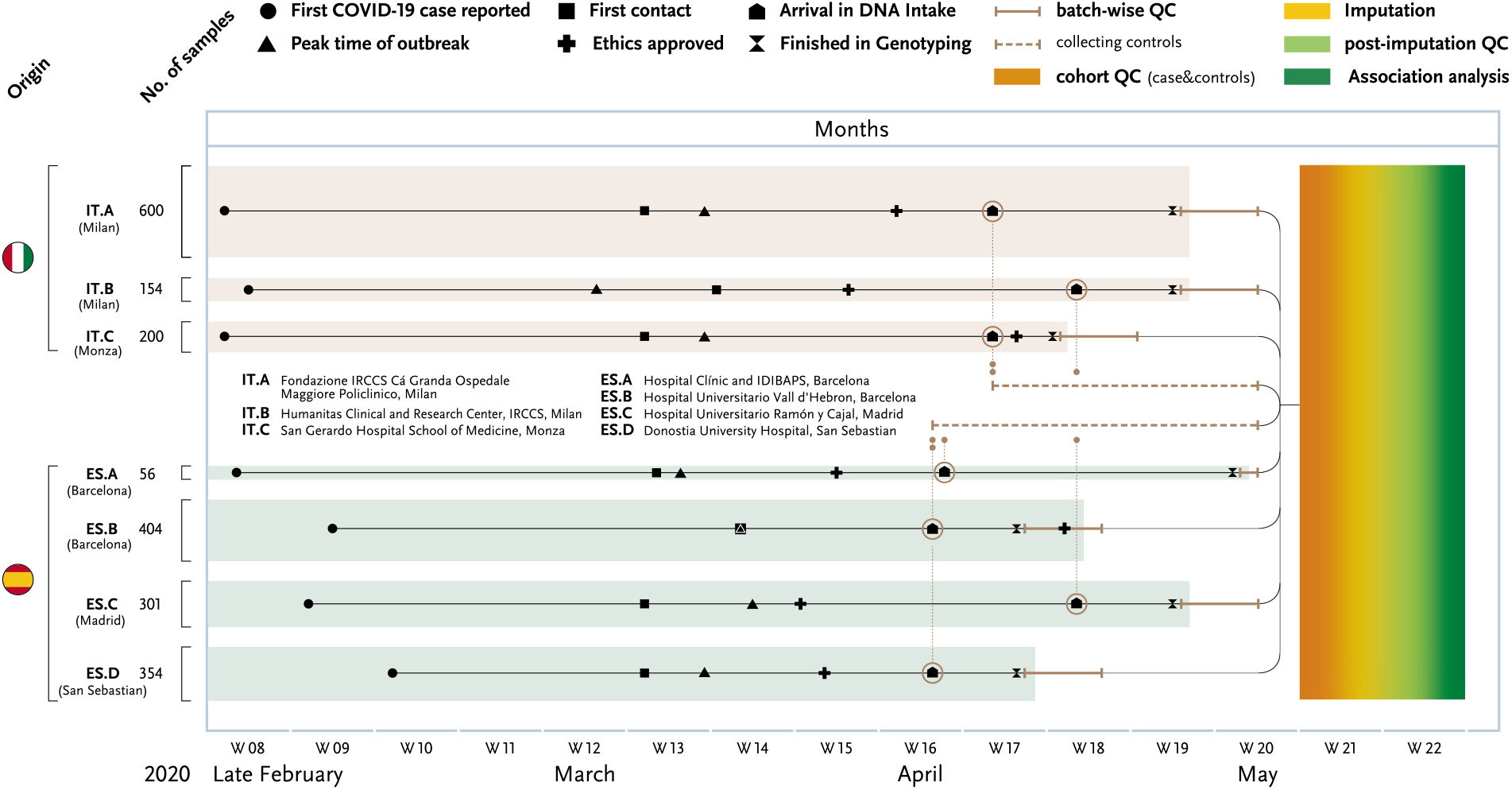
Timeline of rapid Covid-19 genome-wide association study (GWAS). The main events and milestones of the study are summarized in the plot along time (x-axis, weeks (W) are noted). Patient samples from three Italian and four Spanish hospitals were collected around the peak of the local epidemics and ethics applications were quickly obtained through fast-track procedures, i.e. every local ethical review board supported Covid-19 studies by rapid turn-around times, facilitating this fast *de novo* data generation. Within 6 weeks, all collected blood samples were centrally isolated, genotyped and analysed. The rapid workflow from patients to target identification illustrates the utility of GWAS, a standardized tool in research that often relies on international and interdisciplinary cooperation. One centre alone could not have completed this study, not mentioning the increase in statistical power through multi-centre patient contribution. Speed of data production depended heavily on lab automation and speed of analyses reflect existing analytical pipelines and generous support of public so-called “imputation servers” (here, the Michigan imputation server of the Abecasis group).

The pathogenesis of respiratory failure in Covid-19 is poorly understood, but mortality consistently associates with older age and male gender.^6–8^ Clinical associations have also been reported for obesity and cardiovascular disease traits, hypertension and diabetes in particular, but the relative role of these risk factors in determining Covid-19 severity has not yet been clarified.^6–9^ Observations on lymphocytic endothelitis and diffuse microvascular and macrovascular thromboembolic complications may suggest that Covid-19 is a systemic disease that primarily injures the vascular endothelium, but provide mostly hypothetical insights to the underlying pathogenesis in severe Covid-19.^10–12^ On this background, at the peak of the epidemic in Italy and Spain, we performed a genome-wide association study (GWAS) to possibly delineate host genetic factors contributing to respiratory failure in Covid-19. The relatively low Covid-19 disease burden in Norway and Germany allowed for a complementary team to be set up, whereby rapid analysis could occur in parallel with rapid patient recruitment in the affected Italian and Spanish epicenters.

## Materials and methods

### Study subjects / recruitment

We recruited in total 1,980 patients with severe Covid-19 infection defined by hospitalization with respiratory failure and confirmed SARS-CoV-2 viral replication from nasopharyngeal swabs or other relevant biological fluids cross-sectionally from intensive care units and general wards of seven hospitals in five cities in the pandemic epicenters in Italy and Spain (**Table 1** and **Supplementary Table 1A**); Fondazione IRCCS Cá Granda Ospedale Maggiore Policlinico, Milan (n=597), Humanitas Clinical and Research Center, IRCCS, Milan (n=154), San Gerardo Hospital School of Medicine, Monza (n=200), At Hospital Clínic and IDIBAPS, Barcelona (n=56), Hospital Universitario Vall d’Hebron, Barcelona (n=337), Hospital Universitario Ramón y Cajal, Madrid (n=298), Donostia University Hospital, San Sebastian (n=338). Respiratory failure was defined in the simplest possible manner, to ensure feasibility, by requirement of oxygen supplementation or mechanical ventilation, with the severity graded according to maximum respiratory support up until time of blood sampling (oxygen therapy only, non-invasive ventilatory support, invasive ventilatory support, extracorporeal membrane oxygenation). Severity was also binarized to no mechanical oxygenation vs. mechanical oxygenation for severity assessments. Whole blood or buffy coats from diagnostic venipuncture were collected for DNA extraction.

**Table 1.** Overview of patients included in final analysis. Male predominance among patients and the high age with medians > 63 years are consistent among all centres. Sample numbers provided are post-QC for the GWAS (see also **Supplementary Table 1C**).

|  | **Fondazione**<br>**IRCCS Ca'**<br>**Granda**<br>**Ospedale**<br>**Maggiore**<br>**Policlinico,**<br>**Milan**<br>**(IT.A)** | **Humanitas**<br>**Clinical and**<br>**Research**<br>**Center,**<br>**IRCCS,**<br>**Milan**<br>**(IT.B)** | **San**<br>**Gerardo**<br>**Hospital**<br>**School of**<br>**Medicine,**<br>**Monza**<br>**(IT.C)** | **Hospital**<br>**Clínic and**<br>**IDIBAPS,**<br>**Barcelona**<br>**(ES.A)** | **Hospital**<br>**Universitario**<br>**Vall**<br>**d'Hebron,**<br>**Barcelona**<br>**(ES.B)** | **Hospital**<br>**Universitario**<br>**Ramón y**<br>**Cajal,**<br>**Madrid**<br>**(ES.C)** | **Donostia**<br>**University**<br>**Hospital,**<br>**San**<br>**Sebastian**<br>**(ES.D)** |
| --- | --- | --- | --- | --- | --- | --- | --- |
| N | 503 | 140 | 192 | 45 | 228 | 201 | 301 |
| Age, median<br>(IQR) | 64 (54-76) | 67 (57-75) | 66 (56-74) | 69 (59-75) | 65 (56-72) | 69 (60-79) | 67 (57-75) |
| Gender female, n<br>(%) | 159 (32) | 39 (28) | 51 (27) | 13 (29) | 78 (34) | 50 (25) | 124 (41) |
| Respiratory<br>support, n (%) |  |  |  |  |  |  |  |
| - Oxygen only | 0 (0) | 70 (50) | 67 (35) | 7 (16) | 105 (46) | 106 (53) | 255 (85) |
| - Non-invasive<br>ventilation | 399 (79) | 25 (18) | 89 (46) | 6 (13) | 7 (3) | 16 (8) | 0 (0) |
| - Ventilator | 104 (21) | 45 (32) | 33 (17) | 31 (69) | 116 (51) | 77 (38) | 46 (15) |
| - ECMO | 0 (0) | 0 (0) | 3 (2) | 1 (2) | 0 (0) | 2 (1) | 0 (0) |

ECMO: Extracorporeal membrane oxygenation; IQR: interquartile range

For comparison, we included a total of 2,381 controls from Italy and Spain (**Supplementary Table 1B**). We recruited 998 randomly selected blood donors at Fondazione IRCCS Cá Granda Ospedale Maggiore Policlinico, Milan with no evidence of Covid-19 who were genotyped for the purpose of the present study. We also included two control panels with genotype data derived from previous studies using the same genotyping array; from Italy n=396 controls from reference^13^ and from Spain n=987 controls recruited from blood donors (San Sebastian).

### Ethical approval

The project protocol outlined a rapid patient inclusion with principally no additional project-related procedures (material from clinically indicated venipunctures) and with the opportunity of complete anonymity with only minimal data collected. Differences in recruitment and consent procedures between centers were determined by 1) some centers integrating the project in larger Covid-19 biobanking efforts and others doing dedicated inclusion for this project and 2) variability regarding the local ethical committee handling of anonymization vs. deidentification as well as consent procedures. Written informed consent was obtained from all study subjects at each center when possible, alternatively exempt as defined by delayed consent, oral consent or consent via next of kin was collected depending on local ethical committee regulations. For some severely ill patients, where this was not possible, an exemption from informed consent was obtained by the local ethical committee or per local regulations during the Covid-19 pandemic to allow the use of completely anonymized surplus material from diagnostic venipuncture.

The following ethical approvals of the project were obtained from the relevant ethics committees: Germany: Kiel (reference number D464/20); Italy: Fondazione IRCCS Cá Granda Ospedale Maggiore Policlinico (reference number 342_2020 and 334–2020 for cases and controls, respectively), Humanitas Clinical and Research Center, IRCCS (reference number 316/20), San Gerardo Hospital School of Medicine, Monza(the ethics committee of the National Institute of Infectious Diseases Lazzarro Spallanzani reference number 84/2020); Norway: Regional Committee for Medical and Health Research Ethics in South-Eastern Norway (reference number 132550); Spain: Hospital Clínic, Barcelona (reference number HCB/2020/0405), Hospital Universitario Vall d’Hebron, Barcelona (reference PR[AG]244/2020), Hospital Universitario Ramón y Cajal, Madrid (reference number 093/20) and Donostia University Hospital, San Sebastian (reference number PI2020064).

### Sample processing, genotyping and imputation

We performed DNA extraction from all 1,980 cases and 1,394 Italian controls using a Chemagic 360 from PerkinElmer (Waltham, Massachusetts, U.S.) using the low volume kit cmg 1491 and the Buffy coat kit cmg-714 (Chemagen, Baesweiler, Germany), respectively. For genotyping, we used the Illumina’s Global Screening Array (GSA) version 2.0 (GSA v2.0; Illumina Inc., San Diego, USA) that contains 712,189 variants before quality control (QC). Details on genotyping and QC procedures can be found in the **Supplementary Methods**. To maximize genetic coverage, we performed single nucleotide (SNP) imputation on genome build GRCh38 using the Michigan Imputation Server and 194,512 haplotypes generated by the Trans-Omics for Precision Medicine (TOPMed) program (Freeze 5).^14^ After excluding samples during QC (the majority of which were due to population outliers, see **Supplementary Methods** and **Table 1B**) the final case-control datasets comprised 835 patients and 1,255 population derived controls from Italy, and 775 patients and 950 controls from Spain, with a total of 8,965,091 SNPs included for the Italian cohort and 9,140,716 SNPs for the Spanish cohort.

### Statistical methods

To take imputation uncertainty into account, we tested for phenotypic associations with allele dosage data separately for both Italian and Spanish case-control panels through the use of PLINK’s logistic regression framework for dosage data (PLINK v1.9).^15^ Two adjusted association analyses including covariates from principal component analysis (PCA) were conducted (analysis I and II) to control for (I) potential population stratification as well as (II) potential population stratification as well as age and gender bias. A fixed-effects meta-analysis was conducted using the meta-analysis tool METAL^16^ on variants overlapping between both studies using the BETA and its standard error (SE) from the study specific association analyses. We used the commonly accepted threshold of 5 × 10^−8^ for joint P-values to define statistical significance.

Based on results from TOPMed genotype imputation, we utilized three *ABO* SNPs (rs8176747, rs41302905 and rs8176719)^17,18^ to infer ABO blood type and calculated blood group specific odds ratios according to A vs. B/AB/O, B vs. A/AB/O, AB vs. A/B/O and O vs. A/AB/B (see **Supplementary Methods**). To assess in detail the HLA complex at 6p21, at which we found no genome-wide significant associations in the main analysis, we performed sequencing-based HLA typing of 7 HLA loci (*HLA*-*A*, –*C*, –*B*, –*DRB1*, –*DQA1*, –*DQB1*, –*DPB1*) in the subset of 916 cases from Italy and 1,087 cases from Spain (see **Supplementary Methods**), and assessed allelic distribution according to no mechanical ventilation (oxygen supplementation only) vs. mechanical ventilation any type (**Table 1**). A similar assessment was made for lead SNPs rs11385942 and rs657152, and at these broader loci (3p21.31 and 9q34.2) we also performed Bayesian fine-mapping analysis (see **Supplementary Methods**).

## Results

The milestones of the study in the context of the peak outbreaks in Italy and Spain are shown in **Figure 1**. Age, gender and maximum respiratory support up until time of blood sampling for patients included in the final analysis are given in **Table 1** and **Supplementary Table 1**. By utilizing GSA-only data, we were able to perform a uniform quality control of merged Italian and Spanish batches, thus reducing potential batch effects, and conducted Italian and Spanish association analyses (see **Supplementary Methods** and **Supplementary Figure 2** and **3**). Quantile-quantile (Q-Q) plot of the two meta-analyses of Spanish and Italian association results revealed significant associations in the tail of the distribution with minimal genomic inflation (λ_GC_=1.015 and λ_GC_=1.006, respectively; **Supplementary Figure 4**).

We found two loci to be associated with Covid-19 induced respiratory failure with genome-wide significance (P<5×10^−8^) in the meta-analysis (analysis I) (**Figure 2** and **Table 2**), the rs11385942 insertion-deletion GA/A SNP at chromosome 3p21.31, OR_meta_ 1.77 (95% CI, 1.48 to 2.11), P=1.14×10^−10^ and the rs657152 A/C SNP at 9q34.2, OR_meta_ 1.32 (95% CI, 1.20 to 1.47), P=4.95×10^−8^. Both loci showed nominally significant association in both the Spanish and Italian sub-analysis (**Table 2**). Manual inspection of genotype cluster plots of genotyped SNPs in associated regions showed distinct genotype clouds for homozygous and heterozygous calls. Furthermore, an age and gender corrected analysis (analysis II) corroborated observations at both rs11385942 OR_meta_ 2.11 (95% CI, 1.70 to 2.61), P=9.46×10^−12^ and rs657152 OR_meta_ 1.39 (95% CI, 1.22 to 1.59), P=5.35×10^−7^ (**Table 2**; **Supplementary Figure 5**). A further 24 genomic loci showed suggestive evidence (P<1×10^−5^) for association with Covid-19 induced respiratory failure in analysis I (**Supplementary Table 2; Supplementary Figure 6**).

**Table 2.** Susceptibility loci associated with severe Covid-19 with respiratory failure.

| Chr | Association boundaries (bp) | dbSNP id | A1 | A2 | Key genes | Analysis* | Meta-analysis<br>(1610 cases / 2205 controls) |  | Italy<br>(835 cases / 1255 controls) |  |  |  | Spain<br>(775 cases / 950 controls) |  |  |  |
| --- | --- | --- | --- | --- | --- | --- | --- | --- | --- | --- | --- | --- | --- | --- | --- | --- |
|  |  |  |  |  |  |  | P | OR<br>(CI 95%) | P | OR<br>(CI 95%) | A1F<br>cases | A1F<br>ctrs | P | OR<br>(CI 95%) | A1F<br>cases | A1F<br>ctrs |
| 3p21.31 | chr3:45800446-46135604 | rs11385942 | GA | G | *SLC6A20, LZTFL1, FYCO1, CXCR6, XCR1, CCR9* | main | 1.14x10<sup>-10</sup> | 1.77<br>(1.48-2.11) | 1.98x10<sup>-7</sup> | 1.74<br>(1.27-2.38) | 0.14 | 0.09 | 1.32x10<sup>-4</sup> | 1.85<br>(1.50-2.28) | 0.09 | 0.05 |
|  |  |  |  |  |  | corr. age, gender* | 9.46x10<sup>-12</sup> | 2.11<br>(1.70-2.61) | 7.02x10<sup>-8</sup> | 1.95<br>(1.53-2.48) | 0.14 | 0.09 | 1.17x10<sup>-5</sup> | 2.79<br>(1.76-4.42) | 0.09 | 0.05 |
| 9q34.2 | chr9:133257521-133279871 | rs657152 | A | C | *ABO* | main | 4.95x10<sup>-8</sup> | 1.32<br>(1.20-1.47) | 2.90x10<sup>-6</sup> | 1.37<br>(1.20-1.57) | 0.42 | 0.35 | 3.55x10<sup>-3</sup> | 1.26<br>(1.08-1.48) | 0.42 | 0.35 |
|  |  |  |  |  |  | corr. age, gender* | 5.35x10<sup>-7</sup> | 1.39<br>(1.22-1.59) | 5.31x10<sup>-5</sup> | 1.37<br>(1.17-1.60) | 0.42 | 0.35 | 2.81x10<sup>-3</sup> | 1.45<br>(1.13-1.84) | 0.42 | 0.35 |

All association test statistics were adjusted for the top 10 principal components from principal component analysis. \*Two analyses were performed, “main”, only correcting for principal components, and “corr. age, gender”, correcting for age and gender in addition to 10 principal components. In the corrected analysis, 25 controls are excluded from the Spanish and meta-analysis due to missing covariate data. **Chr**: chromosome of marker; **Pos**: Genomic positions were retrieved from NCBI’s dbSNP build v153 (genome build hg38); **Association boundaries**: association boundaries for each index SNP (see **Methods**); **dbSNP id**: rs ID (rs11385942 is annotated as chr3:45834968-45834969:AAA:AA in dbSNPv153 and as chr3:45834967:GA:G in TOPMed imputation reference panel); **A1**: minor and risk allele; **A2**: major allele; **Key gene(s)**: candidate gene(s) in the region; **P/OR**: P-value and corresponding odds ratio and 95% confidence interval with respect to minor allele. For each panel, numbers of cases/controls are displayed in parentheses; **A1F\_cases**: allele frequency of minor/risk allele 1 in cases; **A1F\_ctrs**: allele frequency of minor/risk allele 1 in controls.

**Figure 2.**
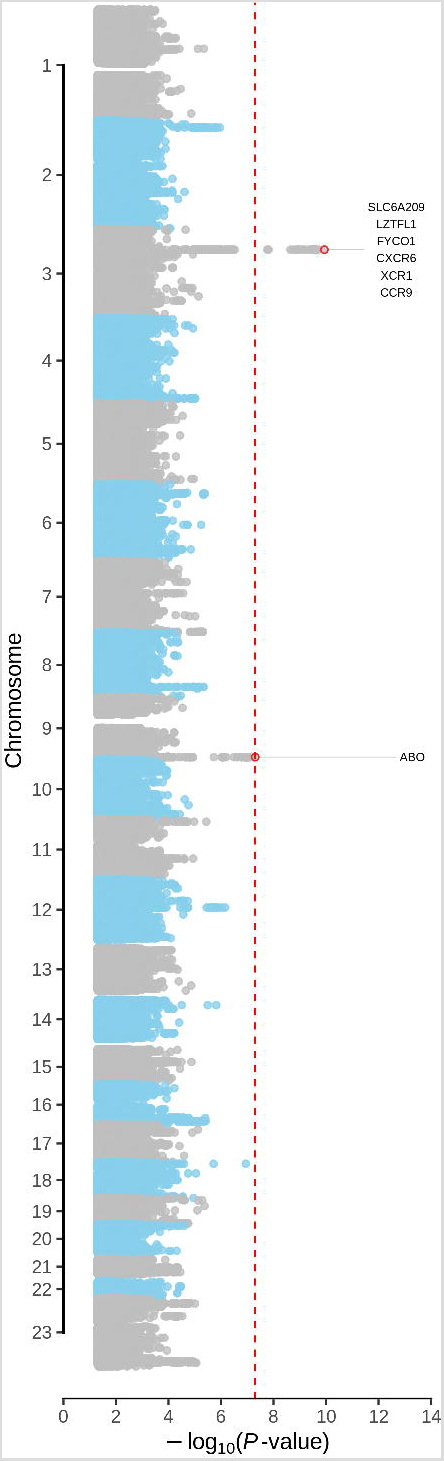
Genome-wide analysis summary (Manhattan) plot of the meta-analysis association statistics highlighting two susceptibility loci with genome-wide significance (P<5×10^−8^) for severe Covid-19 with respiratory failure. Manhattan plot of the association statistics from the meta-analysis controlled for potential population stratification (strategy I). The red horizontal line indicates the genome-wide significance threshold of P=5×10^−8^. **Supplementary Figure 5** shows Manhattan plots including also hits passing a suggestive significance threshold of P<1×10^−5^ (total of 24 additional suggestive genomic loci; see also **Methods** and **Supplementary Table 3**).

Association signals at 3p21.31 and 9q34.2 were fine-mapped to 22 and 38 variants, respectively, with greater than 95% certainty (**Figure 3A** and **3B** and **Supplementary Table 3**). The association signal at 3p21.31 comprised six genes (**Figure 3A** and **Table 2**). We found that the frequency of the risk allele GA of the lead SNP, rs11385942, was higher in patients with mechanical ventilation compared with those receiving oxygen supplementation only with an OR=1.70, 95% CI, 1.27 to 2.26, P=0.00033, in the unadjusted analysis (**Supplementary Table 4**). Available database entries suggest that the frequency of the risk allele of rs11385942 varies between populations world-wide and is monomorphic in China(**Supplementary Figure 7)**.

**Figure 3.**
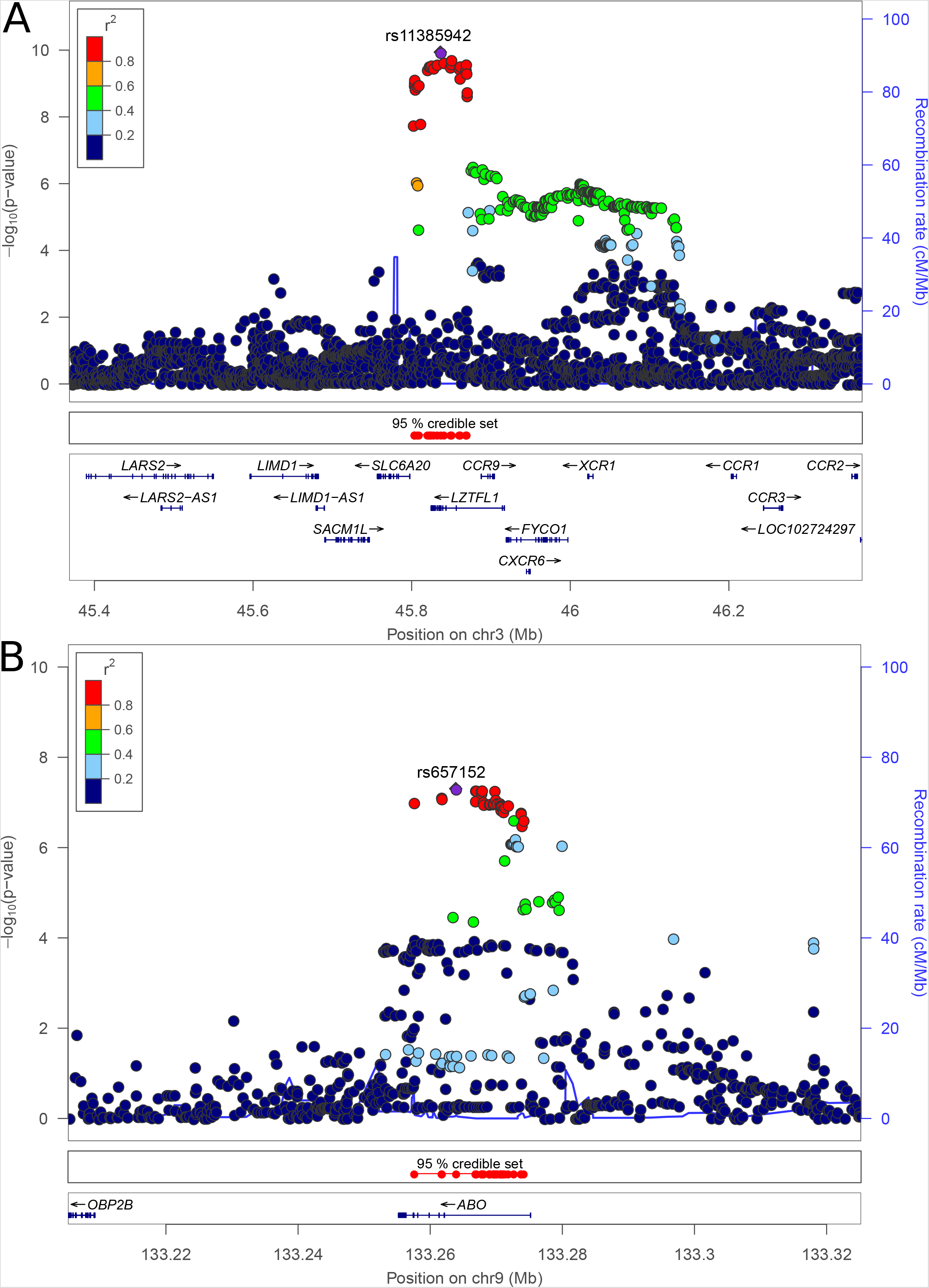
Regional association plots of susceptibility loci associated with severe Covid-19 with respiratory failure. Bayesian fine-mapping analysis (see **Methods)** prioritized 22 and 38 variants for loci 3p21.31 and 9q34.2, respectively, with greater than 95% certainty. LD values were calculated based on genotypes of the merged Italian/Spanish dataset derived from TOPMed imputation (see **Methods**). hg38 positions are plotted.

At 9q34.2 the association signal was restricted to the *ABO* blood group gene (**Figure 3B** and **Supplementary Figure 8**). Accordingly, the distribution of ABO blood groups, as predicted from combinations of genotypes of three different SNPs was skewed in Covid-19 patients with respiratory failure compared with controls (**Supplementary Table 5**), with higher risk for A-positive individuals (meta-analysis result OR=1.45, 95% CI, 1.20 to 1.75, P=1.48×10^−4^) and a protective effect for blood group O (meta-analysis result: OR=0.65, 95% CI, 0.53 to 0.79, P=1.06×10^−5^; see Supplementary Table 5 for details). Both associations and effect directions were consistent in the separate Spain-only and Italian-only case-control analysis (see **Supplementary Table 5**). We found no difference in blood group distribution between patients receiving oxygen supplementation only vs. mechanical ventilation of any kind(**Supplementary Table 5**).

Since several viral infections are known to be controlled by genetic variation at the HLA complex at chromosome 6p21, we scrutinized the extended HLA region (chr6:25–34Mb; **Supplementary Figure 9**). There were no SNP or allele associations signals at the HLA complex meeting neither genome-wide nor suggestive association significance threshold of P=1×10^−5^(**Supplementary Table 6**). Furthermore, we found no significant differences in allelic distribution between patients with oxygen supplementation only and those with mechanical ventilation of any kind (assessed by direct HLA typing, data not shown).

## Discussion

Using a pragmatic approach with simplified inclusion criteria and a complementary team of clinicians at the European Covid-19 epicenters in Italy and Spain and available German and Norwegian scientists, we were able to perform a complete GWAS for Covid-19 respiratory failure in about two months. We detected cross-replicating findings at chromosome 3 and chromosome 9, which achieved genome-wide significance in meta-analysis of both study panels.

On chromosome 3p21 the peak association signal covers a cluster of several genes with potentially relevant functions to severe Covid-19. One notable candidate is *SLC6A20*, which encodes the Sodium/Imino-acid (proline) Transporter 1 (SIT1) that functionally interacts with angiotensin converting enzyme 2 (ACE), the SARS-CoV-2 cell surface receptor.^19,20^ SIT1 expression in the lungs is mainly present in pneumocytes^21^ where SIT1 should be scrutinized for involvement in SARS-CoV-2 viral entry. However, the relevant locus also contains a cluster of genes encoding chemokine receptors, including the CC-motif chemokine receptor 9 (CCR9) and the C-X-C motif chemokine receptor 6 (CXCR6), the latter have been shown to regulate the partitioning of lung-resident memory CD8 T-cells throughout the sustained immune response to airway pathogens, including influenza viruses.^22^ In the publicly available results from the Covid-19 Host Genetics Consortium^23^ a similar association has been observed in an analysis of Covid-19 affected cases vs. a population based sample, however not at genome-wide significant levels, still corroborating our observations. These parallel observations with our analysis, which focused on severe cases with pulmonary failure only, points to the relevance of ascertainment bias in genetic studies of Covid-19, as clinically significant Covid-19 patients are more likely to be included in research projects than asymptomatic cases. The significantly higher frequency of the risk allele at the chromosome 3 locus found in the present study in patients requiring mechanical ventilation compared with oxygen only, provides further support to a role for this genetic region in modifying Covid-19 severity.

Preliminary clinical reports have suggested the involvement of ABO blood groups in Covid-19 susceptibility (preprints by Zhao *et al*.^24^ and Zietz *et al*.^25^). Similar reflections can thus be made for case ascertainment as for the chromosome 3 locus, and ABO blood groups have also been implicated in SARS-CoV-1 susceptibility.^26^ Our data thus aligns with the suggestions that blood group O is associated with lower risk compared with non-O blood groups whereas blood group A is associated with higher risk of acquiring Covid-19 compared with non-A blood groups.^24,25^ Unlike for Chromosome 3, we found no difference between patients receiving oxygen supplementation only and those with mechanical ventilation any kind.^25^ However, it should be noted that the lead SNP at the *ABO* locus in our study (rs657152) has been associated with elevated interleukin-6 (IL-6) levels in childhood obesity in previous GWAS^27^ providing a hypothetical link to the established association of elevated IL-6 with severity and mortality of Covid-19.^28^ Furthermore, genetic variation at the *ABO* locus has previously been associated with a number of procoagulant markers such as von Willebrand factor and Factor VIII, and the potential relationship between our genetic findings and the significant coagulopathy that is observed in severe Covid-19 warrants further attention.

We are fully aware that the pragmatic aspects leading to feasibility of this massive undertaking in a very short period of time during extreme clinical circumstances of the pandemic led to certain limitations that will be important to explore in follow-up studies. For example, to enable recruitment of study participants, a bare minimum of clinical metadata was requested. For this reason, extensive genotype-phenotype elaboration of current findings could not be performed, and adjustments for all potential sources of bias (e.g. underlying cardiovascular and metabolic factors relevant to Covid-19) could not be done. Furthermore, the alignment of our findings with preliminary reports assessing Covid-19 susceptibility should lead to a critical debate as to phenotype definitions for cases and controls in genetic studies of Covid-19. Also, few restrictions during inclusion were made, leading to genotyped samples having to be excluded due to differing ethnicities (genetically population outliers). That said, we took great care to minimize variability between cases and controls arising from such sources, and that could have been introduced from differences between genotyping platforms^29^ e.g. limiting our inclusion of controls to those genotyped on the Illumina Global Screening Array, despite thus reducing our statistical power.

Further exploration of current findings, both as to their utility in clinical risk profiling of Covid-19 patients and mechanistic understanding of the underlying pathophysiology, is now warranted.

## Data Availability

Summary statistics will be made available upon request and after signing of appropriate institutional data transfer agreements, after acceptance of the peer-reviewed manuscript.

## Acknowledgements and funding

The IKMB’s core facilities received infrastructure support by the Deutsche Forschungsgemeinschaft (DFG) Cluster of Excellence “Precision Medicine in Chronic Inflammation” (PMI, EXC2167). The project also received support through a philanthropic donation by Stein Erik Hagen and Canica AS. L.V. was funded by the Fondazione IRCCS Ca’ Granda «COVID-19 Biobank» research grant. We are extremely grateful to all patients who consented to this project and would like to express our condolences to the families of patients passing away due to Covid-19. We further acknowledge the hard and heroic work of the entire clinical staff during the outbreak situation at the different centers and who were able to work on this scientific study in parallel. We thank all Members of the Humanitas COVID-19 Task Force for their contributions to patient recruitment (for full list of members, see Supplementary Material). This work was also supported by the Ministero dell’Istruzione, dell’Università e della Ricerca – MIUR project “Dipartimenti di Eccellenza 2018 – 2022” (n° D15D18000410001) to the Department of Medical Sciences, University of Torino. We thank Fabrizio Bossa and Francesca Tavano for contributing to control sample acquisition and Maria Reig for help in the case sample acquisition. We are thankful to the Basque Biobank (BIOEF, Basque Country, Spain) for assistance in the acquisition of samples. We thank the ethics commissions, review boards and consortia that fast-track reviewed our applications and enabled this rapid genetic discovery study. The IKMB authors received financial support from the UKSH Foundation “Gutes Tun!” (special thanks to Alexander Eck, Jenspeter Horst and Jens Scholz) and the German Federal Ministry of Education and Research (BMBF; grant ID 01KI20197). HLA-Typing was performed and supported by the Stefan-Morsch-Stiftung. M.A.H was supported by the Spanish Ministry of Science and Innovation ‘JdC fellowship IJC2018–035131-I. We would also like to thank Goncalo Abecasis and his team for providing the Michigan imputation server.

## Authorship contributions

T.H.K conceived and initiated the project. T.H.K and A.F. jointly designed, provided infrastructure to and jointly supervised the project. T.H.K, A.F., J.R.H., T.F., D.E. and F.D. wrote the first draft of the manuscript. A.F., D.E. and F.D. coordinated and performed the statistical analyses with contributions from J.K., M.W., L.W., F.U-W. and M.W. S.M., X.Y., A.T., A.P., H.E., M.E.F.B., M.S., N.B., T.W.,W.A., W.P and M.S. performed sample processing, performed DNA extraction and/or genotyping. L.V., L.B., M.B.F., A.A., P.I., R.A., J.F., J.M.B., M.R.G. and M.M.G. organised and supervised patient inclusion and provided intellectual input to study design and the manuscript. M.D.A., S.D., D.P., G.B., C.M., F.A., T.Z., A.B., A.G.C., A.P., A-E.G-F., A.B-G., A.Z., A.B., A.P, A.A., A.I., A.B., A.C-G., A.G., A.L., A.L-F., A.J., A.P., A.V., A.S., B.M., B.N.J., C.Q., C.A., C.C., D.J., D.P., E.S., E.M.P., E.N., F.C., F.M-B., F.P., F.B., F.M., F.R-F., G.G., G.C., G.C., G.F., G.M., H.K., I.M., J.M., J.E., J.A.F., K.G., L.I.S., L.R.B., L.T., L.M., L.S., L.S., L.T., L.R., M.C., M.D.M.R.B, M.G.V., M.H-T., M.A-H., M.D.A., M.B., M.C., M.C., M.C., M.R-G., M.B., M.M., N.M., O.P., P.F., P.P., P.B., P.O., P.T., P.C., P.M.R., R.D.P., R.F.R., R.G, R.N., S.B., S.M., S.A., S.P., S.B., The Humanitas COVID-19 Task Force, T.P., V.R. and V.M. provided samples, phenotypic data and intellectual input to the manuscript. All authors revised and edited the manuscript for critical content and approved of the final version.

## References

1. Zhu N, Zhang D, Wang W, et al. A Novel Coronavirus from Patients with Pneumonia in China, 2019. N Engl J Med 2020;382:727–33.

2. Coronaviridae Study Group of the International Committee on Taxonomy of V. The species Severe acute respiratory syndrome-related coronavirus: classifying 2019-nCoV and naming it SARS-CoV-2. Nat Microbiol 2020;5:536–44.

3. Wu Z, McGoogan JM. Characteristics of and Important Lessons From the Coronavirus Disease 2019 (COVID-19) Outbreak in China: Summary of a Report of 72314 Cases From the Chinese Center for Disease Control and Prevention. JAMA 2020.

4. Berlin DA, Gulick RM, Martinez FJ. Severe Covid-19. N Engl J Med 2020.

5. Marini JJ, Gattinoni L. Management of COVID-19 Respiratory Distress. JAMA 2020.

6. Zhou F, Yu T, Du R, et al. Clinical course and risk factors for mortality of adult inpatients with COVID-19 in Wuhan, China: a retrospective cohort study. Lancet 2020;395:1054–62.

7. Li X, Xu S, Yu M, et al. Risk factors for severity and mortality in adult COVID-19 inpatients in Wuhan. J Allergy Clin Immunol 2020.

8. Mehra MR, Desai SS, Kuy S, Henry TD, Patel AN. Cardiovascular Disease, Drug Therapy, and Mortality in Covid-19. N Engl J Med 2020.

9. Chen R, Liang W, Jiang M, et al. Risk Factors of Fatal Outcome in Hospitalized Subjects With Coronavirus Disease 2019 From a Nationwide Analysis in China. Chest 2020.

10. Levi M, Thachil J, Iba T, Levy JH. Coagulation abnormalities and thrombosis in patients with COVID-19. Lancet Haematol 2020.

11. Varga Z, Flammer AJ, Steiger P, et al. Endothelial cell infection and endotheliitis in COVID-19. Lancet 2020;395:1417–8.

12. Ackermann M, Verleden SE, Kuehnel M, et al. Pulmonary Vascular Endothelialitis, Thrombosis, and Angiogenesis in Covid-19. N Engl J Med 2020.

13. Franke A, McGovern DP, Barrett JC, et al. Genome-wide meta-analysis increases to 71 the number of confirmed Crohn’s disease susceptibility loci. Nat Genet 2010;42:1118–25.

14. Taliun D, Harris DN, Kessler MD, et al. Sequencing of 53,831 diverse genomes from the NHLBI TOPMed Program. bioRxiv 2019:563866.

15. Chang CC, Chow CC, Tellier LC, Vattikuti S, Purcell SM, Lee JJ. Second-generation PLINK: rising to the challenge of larger and richer datasets. Gigascience 2015;4:7.

16. Willer CJ, Li Y, Abecasis GR. METAL: fast and efficient meta-analysis of genomewide association scans. Bioinformatics 2010;26:2190–1.

17. Bugert P, Rink G, Kemp K, Kluter H. Blood Group ABO Genotyping in Paternity Testing. Transfus Med Hemother 2012;39:182–6.

18. Robinson J, Barker DJ, Georgiou X, Cooper MA, Flicek P, Marsh SGE. IPD-IMGT/HLA Database. Nucleic Acids Res 2020;48:D948–D55.

19. Vuille-dit-Bille RN, Camargo SM, Emmenegger L, et al. Human intestine luminal ACE2 and amino acid transporter expression increased by ACE-inhibitors. Amino Acids 2015;47:693–705.

20. Kuba K, Imai Y, Ohto-Nakanishi T, Penninger JM. Trilogy of ACE2: a peptidase in the reninangiotensin system, a SARS receptor, and a partner for amino acid transporters. Pharmacol Ther 2010;128:119–28.

21. Uhlen M, Fagerberg L, Hallstrom BM, et al. Proteomics. Tissue-based map of the human proteome. Science 2015;347:1260419.

22. Wein AN, McMaster SR, Takamura S, et al. CXCR6 regulates localization of tissue-resident memory CD8 T cells to the airways. J Exp Med 2019;216:2748–62.

23. Initiative C-HG. The COVID-19 Host Genetics Initiative, a global initiative to elucidate the role of host genetic factors in susceptibility and severity of the SARS-CoV-2 virus pandemic. Eur J Hum Genet 2020;28:715–8.

24. Zhao J, Yang Y, Huang H, et al. Relationship between the ABO Blood Group and the COVID-19 Susceptibility. medRxiv 2020:2020.03.11.20031096.

25. Zietz M, Tatonetti NP. Testing the association between blood type and COVID-19 infection, intubation, and death. medRxiv 2020:2020.04.08.20058073.

26. Cheng Y, Cheng G, Chui CH, et al. ABO blood group and susceptibility to severe acute respiratory syndrome. JAMA 2005;293:1450–1.

27. Comuzzie AG, Cole SA, Laston SL, et al. Novel genetic loci identified for the pathophysiology of childhood obesity in the Hispanic population. PLoS One 2012;7:e51954.

28. Aziz M, Fatima R, Assaly R. Elevated Interleukin-6 and Severe COVID-19: A Meta-Analysis. J Med Virol 2020.

29. Mitchell BD, Fornage M, McArdle PF, et al. Using previously genotyped controls in genome-wide association studies (GWAS): application to the Stroke Genetics Network (SiGN). Front Genet 2014;5:95.

